# Associations Between Resting State Functional Brain Connectivity and Childhood Anhedonia: A Reproduction and Replication Study

**DOI:** 10.1101/2022.10.24.22281441

**Authors:** Yi Zhou, Narun Pat, Michael C. Neale

## Abstract

**Background:** Previously, a study using a sample of the Adolescent Brain Cognitive Development (ABCD)® study from the earlier 1.0 release found differences in several resting state functional MRI (rsfMRI) brain connectivity measures associated with children reporting anhedonia. Here, we aim to reproduce, replicate, and extend the previous findings using data from the later ABCD study 4.0 release, which includes a significantly larger sample.

**Methods:** To reproduce and replicate the previous authors’ findings, we analyzed data from the ABCD 1.0 release (n = 2437), in an independent subsample from the newer ABCD 4.0 release (n = 6456), and in the full ABCD 4.0 release sample (n = 8866). Additionally, we assessed whether using a multiple linear regression approach could improve replicability by controlling for the effects of comorbid psychiatric conditions and socio-demographic covariates.

**Results:** We could only replicate the significant association between anhedonia and the *Within Cingulo-Opercular network connectivity* measure in an independent subsample of the ABCD 4.0 data release. When using the larger full ABCD 4.0 sample, six out of the eleven previously reported associations remained significant. Accounting for socio-demographic covariates and comorbid conditions using multiple linear regression did not improve replicability but allowed for the identification of specific and independent effects of anhedonia on 16 rsfMRI connectivity measures in the full ABCD 4.0 release sample.

**Conclusion:** Replication of previous findings were limited. A multiple linear regression approach helped resolve the specificity of rsfMRI connectivity associations with anhedonia.

## Introduction

Anhedonia is defined as a markedly diminished interest or pleasure in previously enjoyable activities and is a transdiagnostic symptom that is a core component of major depressive disorders (MDD) (1) and schizophrenia (SZN) (2). Symptoms of anhedonia are also present in substance use disorders (3), PTSD (4), bipolar depression (Gałuszko-Węgielnik et al., 2019), and ADHD (5). Anhedonia in children and adolescents is a significant prognostic predictor of greater depression severity (6), treatment resistant depression (7), and suicidal behaviors (8).

Functional neuroimaging approaches have been widely used to explore the neurocircuitry of anhedonia (9). Functional brain connectivity is a measure of the coactivation of different brain regions, which measures the degree of synchrony between the blood oxygen level dependent (BOLD) signals across time between regions in the brain (Lv et al., 2018). In other words, functional connectivity allows for the characterization of networks of brain activity rather than activity in single brain regions. Importantly, functional connectivity can be measured at rest.

While there have been many studies of brain activity and connectivity in anhedonic adults, fewer have been conducted in children and adolescents. However, findings from these studies generally converge on the significance of disruptions in the reward, default mode, and salience networks (10–12). A recent study using the early 1.0 release of the Adolescent Brain Cognitive Development (ABCD) study data found several resting state functional MRI (rsfMRI) brain network connectivity measures associated with anhedonia in children aged 9-10 years old (13). Importantly, it was one of the largest studies of anhedonia in children with a sample size of ∼2,500 participants including 215 children reporting past and/or present anhedonia. Presently, the latest 4.0 release of the ABCD study, which includes a significantly larger sample of participants with neuroimaging and behavioral data (n = 11,878), has been made available. Given that prior releases of the ABCD data are archived and publicly available, there is a significant opportunity to both *reproduce* and *replicate* these findings.

We define *reproducibility* as the ability to achieve exactly the same results as a previous study by using the same data and analytical approach, and *replicability* as the ability to achieve the same (or similar) results as a previous study in a different dataset (14). By our definition, reproducibility is better able to assess the consistency of results while replicability is better able to assess the generalizability of those results. The aims of this present study are to reproduce the previously reported associations between rsfMRI connectivity and childhood anhedonia using the ABCD 1.0 release sample, and to replicate those findings using an independent subset of the larger ABCD 4.0 release sample, excluding participants from the ABCD 1.0 release sample.

Importantly, in depressive disorders, anhedonia is characterized as the loss of pleasure and interest that is distinct from feelings of sadness or other dysphoric moods(15). Thus, there is great need to elucidate the specific neurobiological underpinnings associated with anhedonia, distinct from other comorbid symptoms, to better understand the underlying brain dysfunction. Thus, we also aim to extend our analyses and evaluate the specificity of rsfMRI connectivity associations with anhedonia by evaluating the effects of significantly comorbid psychiatric symptoms and diagnoses.

## Methods and Materials

All of our analyses were performed in *R (version 4*.*0*.*3)* and *Rstudio*. We used several scripts, including the *utils*.*R* and *combat*.*R*(16) scripts for data harmonization, which were provided by the previous authors immediately upon request. The code and data structures used for our study can be accessed from the associated NDA study.

The code we used can also be found at our Open Science Framework repository (https://osf.io/vy85h/?view_only=0497385708874a6a9cce2bbfc5c30600).

### ABCD Study Data

The ABCD® study is the largest longitudinal study of brain development in children in the United States (https://abcdstudy.org/). The study has collected structural and functional brain imaging measures as well as detailed psychiatric and behavioral data from almost 12,000 children starting from when they were 9-10 years old. Notably, data is released on a continuous basis. For this study, we used baseline data from the ABCD 1.0 and ABCD 4.0 releases.

#### rsfMRI Connectivity Measures and Quality Control (QC)

Neuroimaging processing pipelines and analyses for the ABCD study are reviewed by (17). Briefly, the functional scans include twenty minutes of resting-state data acquired with eyes open and passive viewing of a crosshair (18). From the ABCD Data Repository, we obtained rsfMRI connectivity measures which were constructed using a seed-based correlational approach where regions of interest (ROIs) within Gordon parcellations (19) were grouped together into predefined cortical networks.

Briefly, correlations between unique pairs of ROI’s were obtained and Fisher transformed into z-statistics. Connectivity measures represent the averaged Fisher-transformed correlations of all the unique pairs of ROIs either within a cortical network, between cortical networks, or between cortical networks and subcortical regions. From the ABCD Data Repository, we obtained rsfMRI connectivity measures between 19 subcortical regions and 12 cortical networks (data structure: mrirscor02), as well as rsfMRI connectivity measures from within and between the 12 cortical networks (data structure: abcd_betnet02). Thus, there were 228 (12 × 19) subcortical ROI vs. cortical network rsfMRI connectivity measures and 78 (12**C**2 network pairs + 12 within network) within/between cortical network rsfMRI variables, for a total of 306 rsfMRI connectivity measures.

For QC, we used the IQC_RSFMRI_GOOD_SER variable, which represents the number of rsfMRI runs that were complete, passed protocol compliance and QC, and had field maps acquired within 2 scans prior to the run that were complete and passed QC and protocol compliance. Like the previous authors’, we retained subjects who had IQC_RSFMRI_GOOD_SER values greater than or equal to four.

For analyses using only ABCD 1.0 release sample, we also removed individuals who were scanned by “Philips Medical Systems” MRI machines because of a post-processing issue in the ABCD 1.0 release, which was resolved in later releases. When working with ABCD 4.0 release sample, we retained all the subjects who were scanned by *Philips Medical Systems* scanners because the post-processing errors identified in the ABCD 1.0 release had been fixed for the ABCD 4.0 release.

#### Data Harmonization

Different MRI scanners were used across the 21 sites in the ABCD study. The original authors harmonized the data across MRI scanners by using the ComBat tool (*combat*.*R)* to adjust for batch effects due to the different scanners used. Here, we do the same and harmonize the data separately for the subcortical ROI vs. cortical network and within/between cortical network rsfMRI measures as it is possible these two variable types may be affected by scanners differently(20). Note, before the harmonization step, listwise deletion of subjects with any missing rsfMRI data was done as data harmonization requires complete data. There were 23 different scanners used in the study for the ABCD 1.0 release and 29 different scanners for the ABCD 4.0 release. Thus 23 and 29 batch effects were used to adjust the ABCD 1.0 and 4.0 releases, respectively.

#### Psychiatric Symptoms and Diagnoses

The focus of this study were past/present symptoms of anhedonia. However, we were also interested in other psychiatric conditions that may be comorbid with anhedonia. Psychiatric data were obtained from the youth (*data structure: abcd_ksad501)* and parent *(data structure: abcd_ksad01)* Kiddie Schedule for Affective Disorders and Schizophrenia (KSADS) data structures from the ABCD study. Using both youth and parent KSADS items, we combined past and present items for the same symptom or diagnosis and consolidated some items into a single variable. For example, we consolidated 18 past and present suicide related diagnosis items into one single suicide thoughts and behavior variable, which was similarly done in another study (21).

For psychiatric conditions besides anhedonia, we first selected the KSADS items representing psychiatric diagnoses and not individual symptoms. However, in both the Youth and Parents KSADS data, no diagnosis variables for major depressive disorder (MDD) were available. Thus, we selected two MDD related symptoms (besides anhedonia), irritability and depressed mood, from both the Youth and Parent data to be used in our analyses. Similarly, no diagnostic variable was available for ADHD. Thus, we created a representative variable, inattention_distracted_p, which is a combination of two prevalent ADHD related symptom items (*Symptom - Difficulty sustaining attention since elementary school* and/or *Symptom - Easily distracted since elementary school*) from the parent KSADS data.

### Statistical Analyses

#### Student’s and Bayes Factor T-Tests

Prior to statistical analyses, participants with missing data for psychiatric symptoms/diagnoses were removed. We then proceeded to identify and remove outliers for each rsfMRI connectivity measure as values 1.5 times greater than the interquartile range (IQR) of values.

We performed Student’s T-Tests for each of the 306 rsfMRI measures between controls and individuals with past and/or present psychiatric symptoms/diagnoses of interest (the reference group consisted of individuals endorsing psychiatric symptoms/diagnoses, such as anhedonia). We applied the Benjamini-Hochberg adjustment for multiple testing corrections. Finally, we performed Bayes Factor T-Tests for each of the rsfMRI measures and reported natural logarithms of the Bayes Factors (lnBF). As with the previous authors’, lnBF values greater than 1.1 were considered significant.

#### Tetrachoric Correlations

Tetrachoric correlations are suitable for use with binary or categorical variables as it assumes responses arise from an underlying normal distribution with thresholds that delineate response categories. The *tetrachoric()* function from the *psych* R package was used in our analyses. Tetrachoric correlations were used to estimate the correlations between pairs of 34 binary psychiatric variables.

#### Multiple Linear Regressions

For multiple linear regression analyses, the ABCD rsfMRI data (from both 1.0 and 4.0 releases) were harmonized for MRI scanner using the ComBat tool as previously described, except we also adjusted for batch effects with covariates (20). The covariates included during data harmonization were: age, sex, race/ethnicity, anhedonia, bipolar II, irritability, and depressed mood.

We performed linear mixed effects modeling using the *lmer4* package in *R*, a form of multiple linear regression, on the harmonized data. Individual rsfMRI connectivity measures were modeled as outcome variables while age, sex, race/ethnicity, anhedonia (reference group was the control group), depressed mood, irritability, and bipolar II disorder were modeled as independent explanatory variables. Age, sex, and race/ethnicity were considered potential confounding variables. The independent effects of anhedonia, bipolar II, irritability, and depressed mood symptoms on rsfMRI connectivity measures were assessed by identifying corresponding statistically significant partial regression coefficients, after multiple testing corrections. Family ID was included as a random effect to control for the non-independence of values from participants who belonged to the same family. We adjusted for multiple comparisons with the Benjamini-Hochberg method to control for a False Discovery Rate of 0.05.

## Results

### Reproduction of previous findings

To reproduce the previous authors’ results, we used the ABCD 1.0 release sample. Socio-demographic characteristics for this sample can be found in Table 1. Note that although the two groups appear to exhibit differences in a few of these characteristics, they were not controlled for statistically when we performed in our t-tests in order to remain consistent with the previous authors’ approach.

**Table 1.**
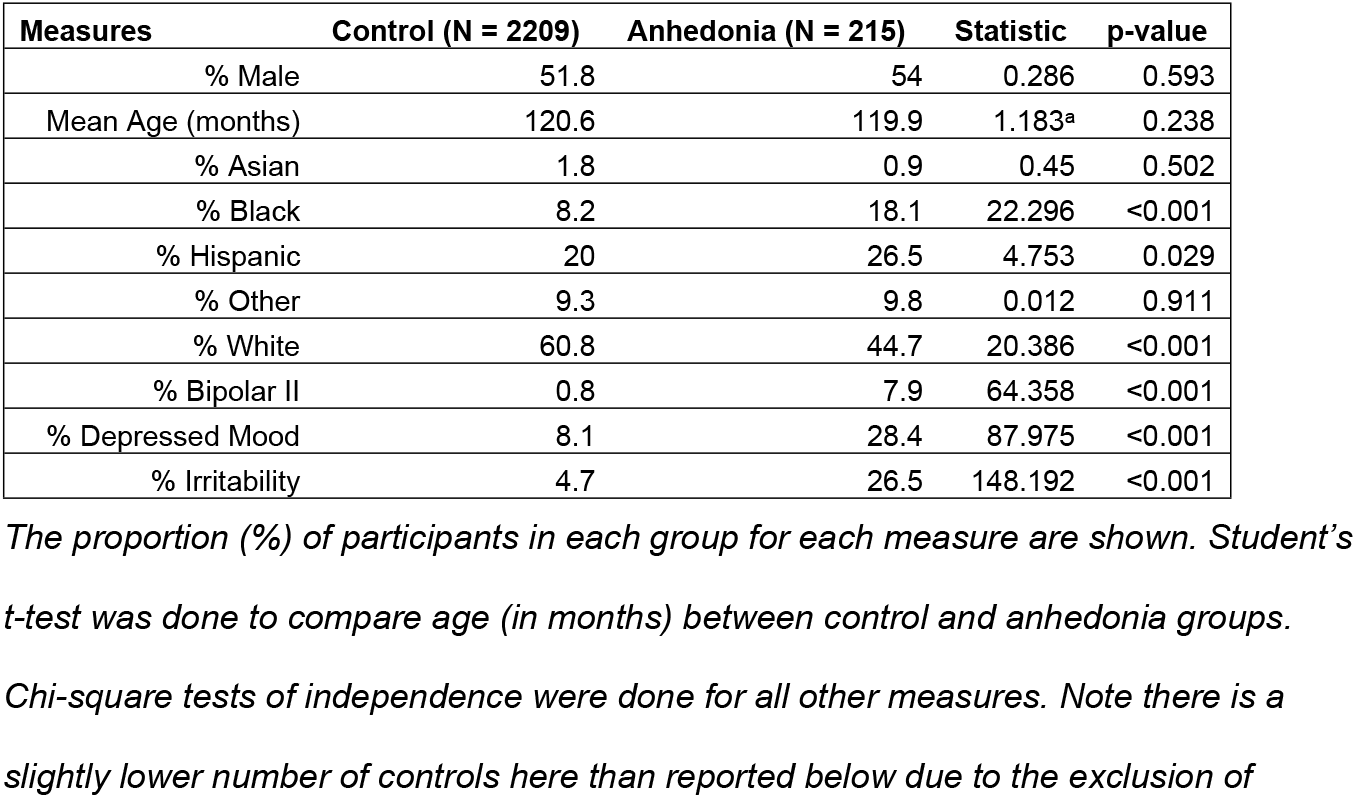
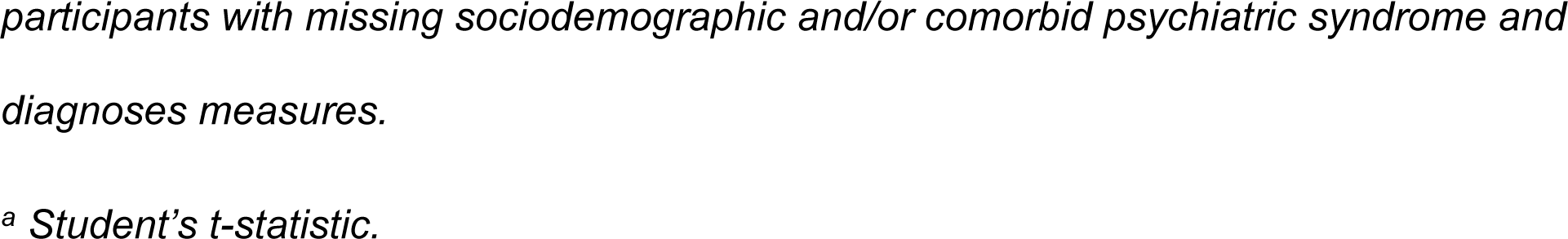
Comparison of Socio-demographic Measures Between Controls and Those with Anhedonia in the ABCD 1.0 Release Sample.

We were able to successfully reproduce their findings (13). To assess how well we reproduced the previous authors’ results, we calculated Pearson correlations of the t-statistics and lnBF values between our values and those reported by the previous authors when comparing individuals with or without anhedonia (Fig. 1A and 1B).

**Figure 1.**
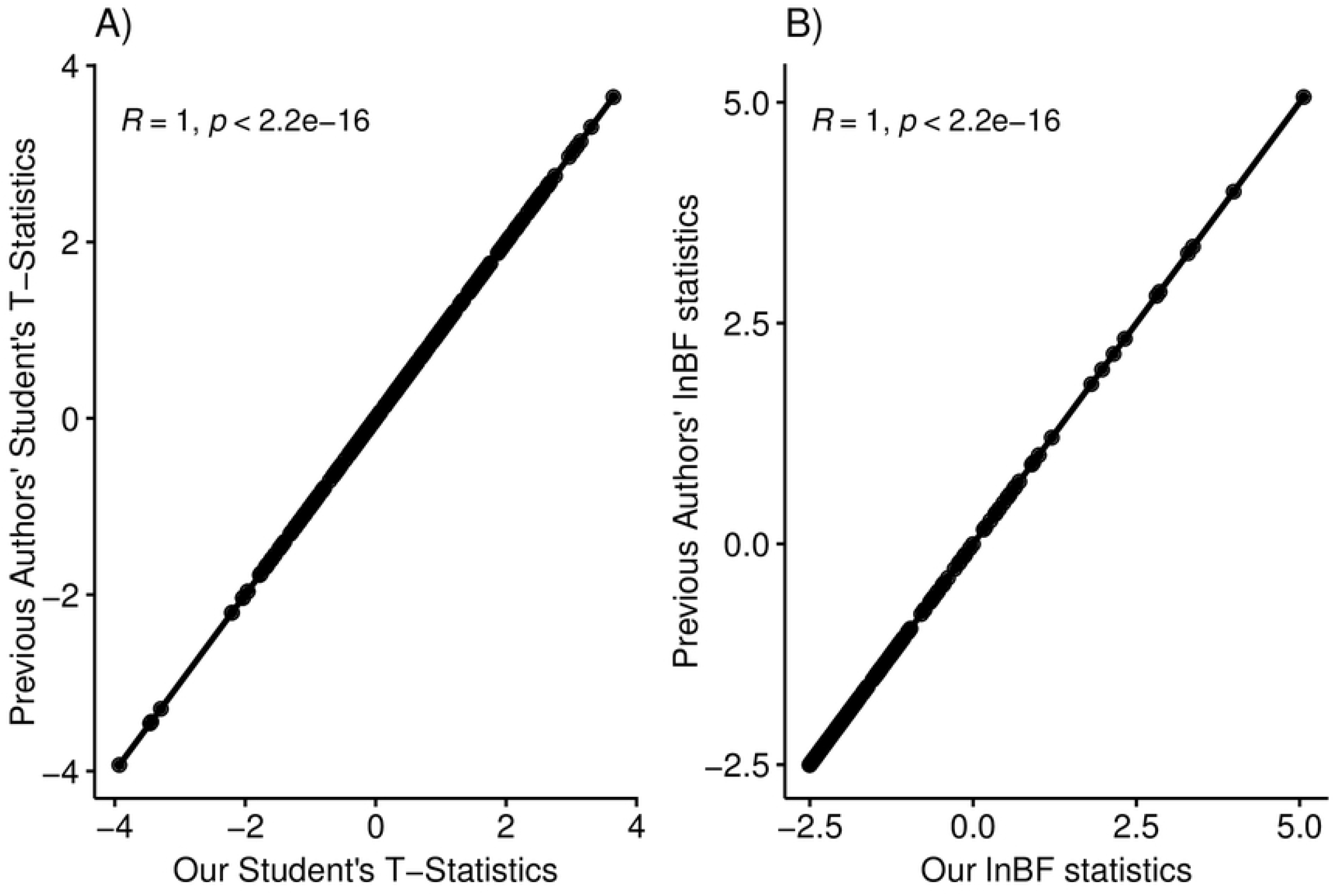
Reproduction of previous statistical analyses. Pearson correlations between A) Student’s t-statistics and B) lnBF statistics derived from the previous author’s analyses and those derived from our replication analyses. LnBF – natural logarithm of Bayes Factors.

Like the previous authors, we identified 215 individuals who endorsed past and/or present anhedonia and 2,222 controls who reported neither past nor present anhedonia at the baseline timepoint. In line with the previous authors findings, 11 rsfMRI connectivity measures were found to be associated with anhedonia using the lnBF(10) statistic (Table. 2; full table of results found in Supplementary Table. 1). To be consistent with the previous authors, individuals with anhedonia were the reference group.

**Table 2.**
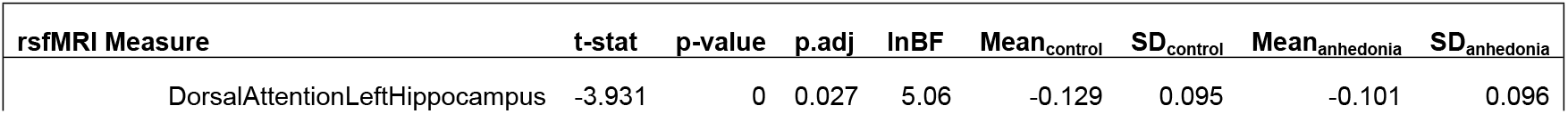
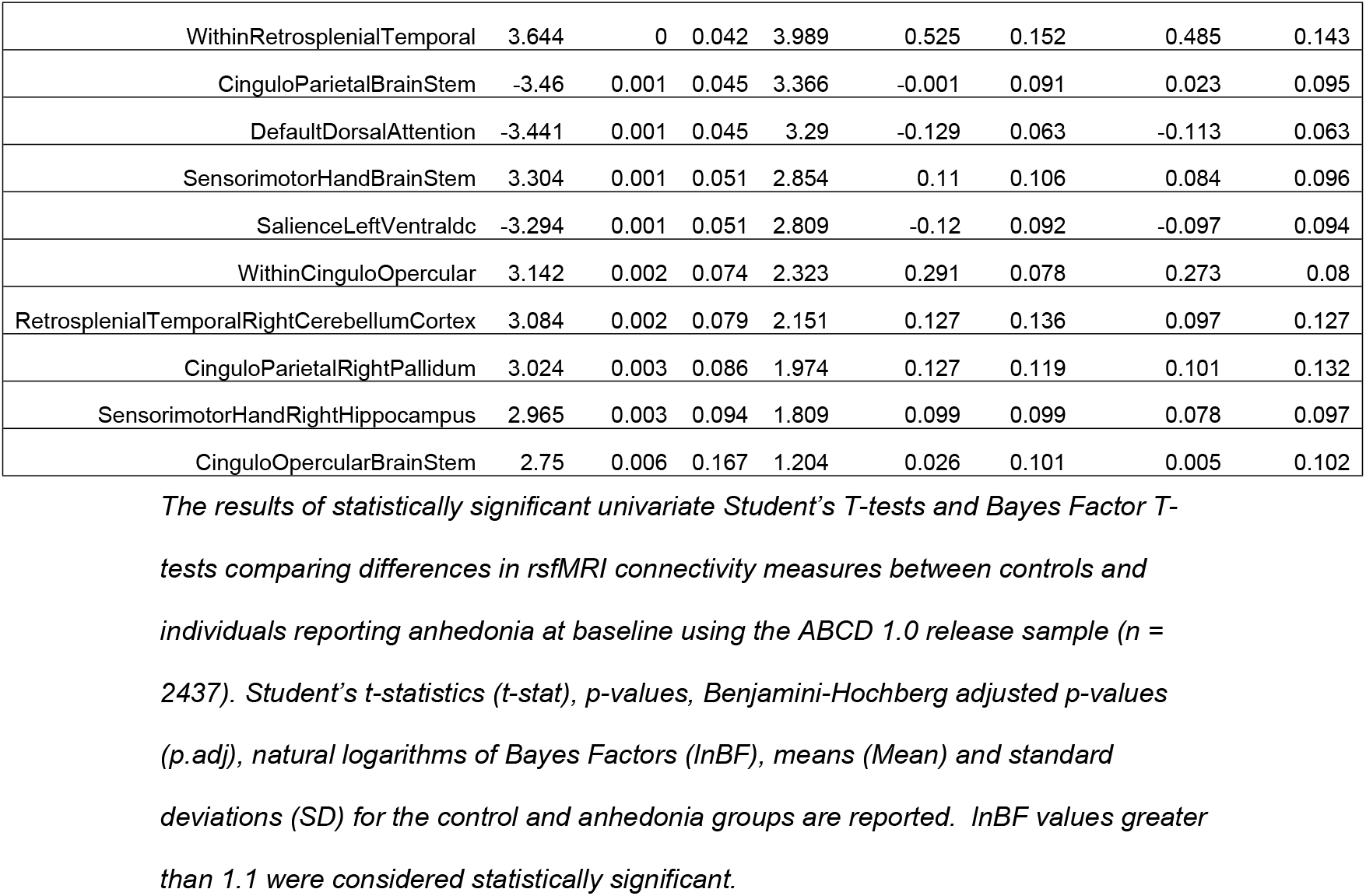
Reproducing Results of rsfMRI Network Connectivity Measures Associated with Anhedonia.

### Replication of previous findings

At the time of writing, the ABCD 4.0 release has been made available. We wanted to *replicate* the previous findings by using the full cohort, excluding the subjects used in the previous analyses, which is analogous to replication in an independent sample. In this sub-sample of the ABCD 4.0 release (which excludes participants from ABCD 1.0 release), we found 591 participants who endorsed past and/or present anhedonia and 5,865 controls who did not. Socio-demographic characteristics for this sample can be found in Table 3.

**Table 3.** Comparison of Sociodemographic Measures Between Controls and Those with Anhedonia in the ABCD 4.0 Release, Excluding ABCD 1.0 Release, Sub-sample.

| Measures | Control (N = 5863) | Anhedonia (N = 591) | Statistic | p-value |
| --- | --- | --- | --- | --- |
| % Male | 51.2 | 56.7 | 6.202 | 0.013 |
| Mean Age (months)<sup>a</sup> | 118.5 | 119.1 | -1.705 | 0.089 |
| % Asian | 2.3 | 0.8 | 4.703 | 0.03 |
| % Black | 15.2 | 20.8 | 12.449 | <0.001 |
| % Hispanic | 20.5 | 26.4 | 11.087 | 0.001 |
| % Other | 10.4 | 12.9 | 3.055 | 0.08 |
| % White | 51.6 | 39.1 | 33.287 | <0.001 |
| % Bipolar II | 0.3 | 8.3 | 313.34 | <0.001 |
| % Depressed Mood | 6.4 | 31.8 | 431.052 | <0.001 |
| % Irritability | 4.4 | 27.9 | 484.563 | <0.001 |

*The proportion (%) of participants in each group for each measure are shown. Student's t-test was done to compare age (in months) between control and anhedonia groups. Chi-square tests of independence were done for all other measures. Note there is a slightly lower number of controls here than reported above due to the exclusion of participants with missing sociodemographic and/or comorbid psychiatric syndrome and diagnoses measures.*
<sup>a</sup> Student's t-statistic.

When comparing the groups using Student’s and Bayes Factor t-tests, 18 rsfMRI connectivity measures were found to be significantly associated with anhedonia according to the lnBF statistic (Table. 4; full table of results found in Supplementary Table 2). However, only the w*ithinCinguloOpercular network* rsfMRI connectivity identified by the previous authors, was also found to be significant.

**Table 4.**
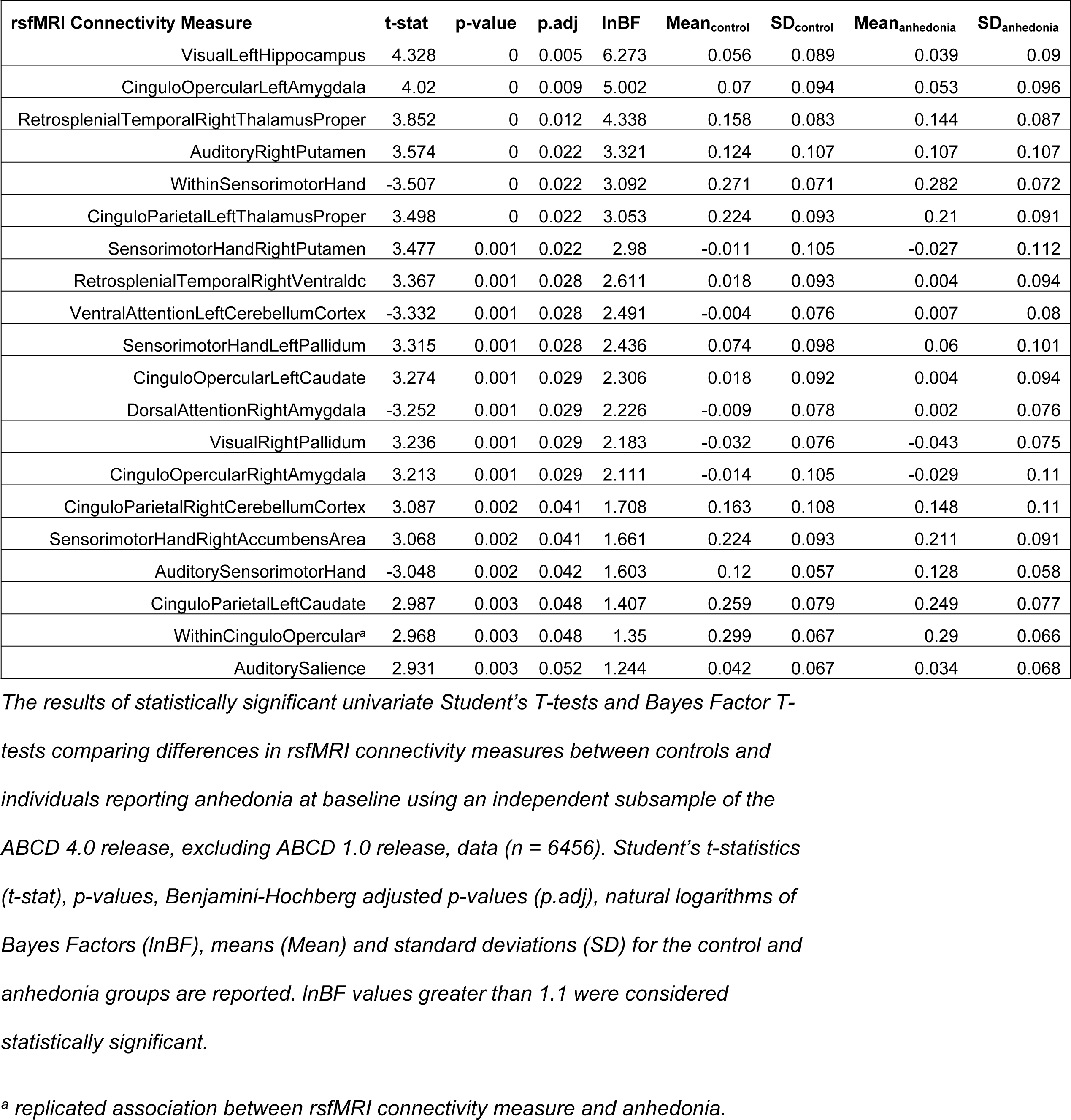
Replicating Results of rsfMRI Connectivity Measures Associated with Anhedonia.

To increase our power to detect genuine associations, we next performed our analyses on the full ABCD 4.0 release sample, including participants from the ABCD 1.0 release. In the full ABCD 4.0 release sample, there were 800 participants who endorsed past and/or present anhedonia and 8066 controls who did not. Socio-demographic characteristics for this sample can be found in Table 5.

**Table 5.** Comparison of Sociodemographic Measures Between Controls and Those with Anhedonia in the Full ABCD 4.0 Release Sample.

| Measures | Control (N = 8064) | Anhedonia (N = 800) | Statistic | p-value |
| --- | --- | --- | --- | --- |
| % Male<sup>a</sup> | 51.3 | 55.8 | 5.589 | 0.018 |
| Mean Age (months) | 119.1 | 119.3 | -0.845 | 0.398 |
| % Asian<sup>a</sup> | 2.2 | 0.9 | 5.363 | 0.021 |
| % Black<sup>a</sup> | 13.3 | 19.8 | 25.19 | <0.001 |
| % Hispanic<sup>a</sup> | 20.4 | 26.6 | 16.579 | <0.001 |
| % Other | 10.1 | 12.1 | 2.916 | 0.088 |
| % White<sup>a</sup> | 54 | 40.6 | 52.043 | <0.001 |
| % Bipolar II<sup>a</sup> | 0.5 | 7.9 | 347.62 | <0.001 |
| % Depressed Mood<sup>a</sup> | 6.9 | 30.9 | 507.29 | <0.001 |
| % Irritability<sup>a</sup> | 4.4 | 27.4 | 627.02 | <0.001 |

*The proportion (%) of participants in each group for each measure are shown. Student's t-test was done to compare age (in weeks) between control and anhedonia groups. Chi-square tests of independence were done for all other measures. Note there is a slightly lower number of controls here than reported below due to the exclusion of participants with missing sociodemographic and/or comorbid psychiatric syndrome and diagnoses measures.*
<sup>a</sup> Student's t-statistic.

Notably, 6 out of the 11 rsfMRI connectivity measures identified by the previous authors were also significantly associated with anhedonia in this analysis (Table. 6; full table of results found in Supplementary Table 3). As well, 17 out of the 20 significant rsfMRI connectivity measures identified in the independent sub-sample of the ABCD 4.0 release (from Table. 4) were also found to be significant in the analyses using the full ABCD 4.0 release sample, including the *within CinguloOpercular* network connectivity measure (Table. 6).

**Table 6.**
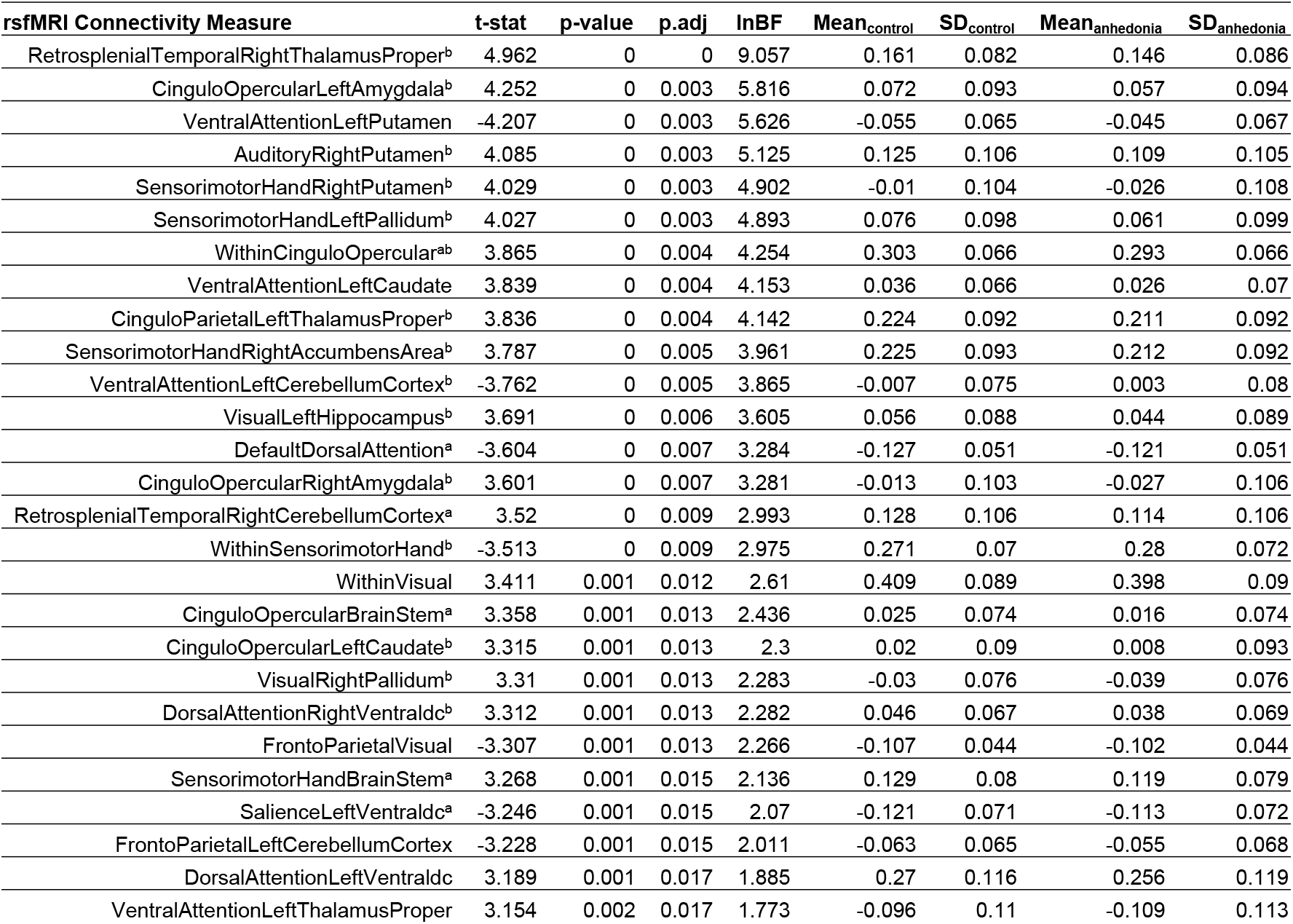
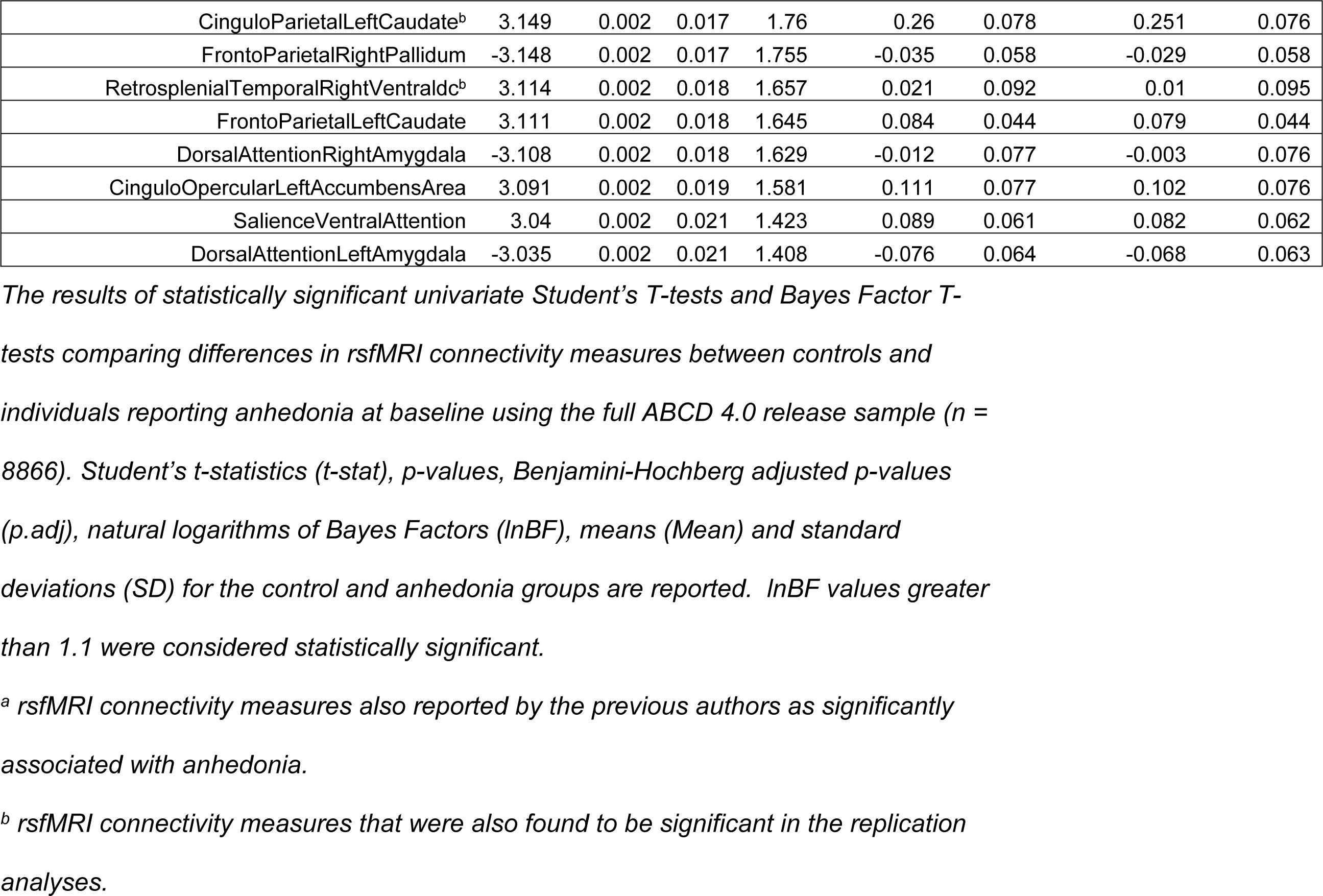
Significant Associations Between rsfMRI Connectivity Measures and Anhedonia in the Full ABCD 4.0 Sample.

It is important to note that individuals reporting anhedonia may also report other symptoms or psychiatric diagnoses. Thus, it is important to evaluate the specificity of the associations between rsfMRI connectivity and anhedonia. In order to identify psychiatric conditions significantly comorbid in individuals reporting anhedonia, we performed tetrachoric correlations between anhedonia and 33 additional psychiatric diagnoses and symptoms collected at baseline (Supplementary Figure. 1). Three psychiatric conditions exhibited correlation coefficients greater than or equal to 0.5 with anhedonia: irritability, depressed mood, and bipolar II disorder. In the full ABCD 4.0 release sample, 32%, 28%, and 8% of the participants reporting anhedonia also reported depressed mood, irritability, and bipolar II disorder (Supplementary Table. 4).

We next performed student’s and Bayes Factor t-tests to compare rsfMRI connectivity between individuals with and without depressed mood, irritability, and bipolar II, respectively (Supplementary Tables. 5-7). We found that several rsfMRI connectivity measures associated with anhedonia were also associated with these other psychiatric conditions.

### Multiple Linear Regression Approach

While we were able to characterize which rsfMRI connectivity measures were specifically associated with anhedonia vs. shared with other psychiatric conditions, simple t-tests were not able to estimate the independent effects of each psychiatric condition on rsfMRI connectivity. Furthermore, t-tests are not able to control for potentially confounding sociodemographic variables such age, sex, and race/ethnicity. Chi-square tests of independence showed significant differences in race/ethnicity and sex between individuals reporting anhedonia and those who do not, across the different samples used in our analyses (Tables 1, 3, and 5).

By using a multiple linear regression approach where a rsfMRI connectivity measure is modeled as the response (or outcome) variable, comorbid psychiatric conditions as well as confounding factors can be included as explanatory (or predictor) variables. Thus, multiple linear regression allows for the estimation of the main effects of anhedonia on rsfMRI connectivity, independent of the effects of depressed mood, irritability, and bipolar II disorder (and vice versa), and the effects of confounding covariates.

Since the ABCD 1.0 release sample and the subsample of ABCD 4.0 release, excluding the ABCD 1.0 sample, were not matched on age, sex, or race/ethnicity, we hypothesized that controlling for these potential confounders may improve replicability. Thus, we first performed multiple linear regression in the ABCD 1.0 release sample and then in the ABCD 4.0 release sample, excluding the ABCD 1.0 release sample. The reference group was the control group without symptoms with anhedonia. In the ABCD 1.0 release sample, only two rsfMRI connectivity measures exhibited statistically significant partial regression coefficients for anhedonia, after multiple testing corrections (Table 4; full multiple regression results found in Supplementary Table 8).

**Table 4.** rsfMRI Connectivity Measures with Significant Partial Regression Coefficients for Anhedonia in the ABCD 1.0 Sample.

| rsfMRI Connectivity Measure | Effect of Anhedonia | Std.Err | t-value | p-value | p.adj | R<sup>2</sup> |
| --- | --- | --- | --- | --- | --- | --- |
| DorsalAttentionLeftHippocampus | 0.024 | 0.007 | 3.208 | 0.001 | 0.019 | 0.012 |
| CinguloParietalBrainStem | 0.021 | 0.007 | 2.968 | 0.003 | 0.036 | 0.006 |

*Statistically significant effects of anhedonia as a predictor of rsfMRI connectivity measures from multiple linear regression tests are shown. The standard errors (Std.Err), t-values, p-values, Benjamini-Hochberg adjusted p-values (p.adj) for the effects of anhedonia, and overall coefficients of determination (R<sup>2</sup>) for each regression are reported.*

It is likely that including the additional explanatory variables into the model reduced the statistical power to detect significant associations in the relatively small sample. Accordingly, while more rsfMRI connectivity measures were significantly associated with anhedonia in the ABCD 4.0 release, excluding ABCD 1.0 release, sub-sample, we were unable to replicate the associations found using the ABCD 1.0 release sample (Table. 5). rsfMRI connectivity measures with significant partial regression coefficients for bipolar II and depressed mood were also identified for analyses in the ABCD 1.0 release sample (Supplementary Table. 8) and for depressed mood, irritability, and bipolar II in the ABCD 4.0 release, excluding ABCD 1.0 release, sub-sample (Supplementary Table. 9). No significant partial regression coefficients for irritability were detected in the ABCD 1.0 release sample.

**Table 5.**
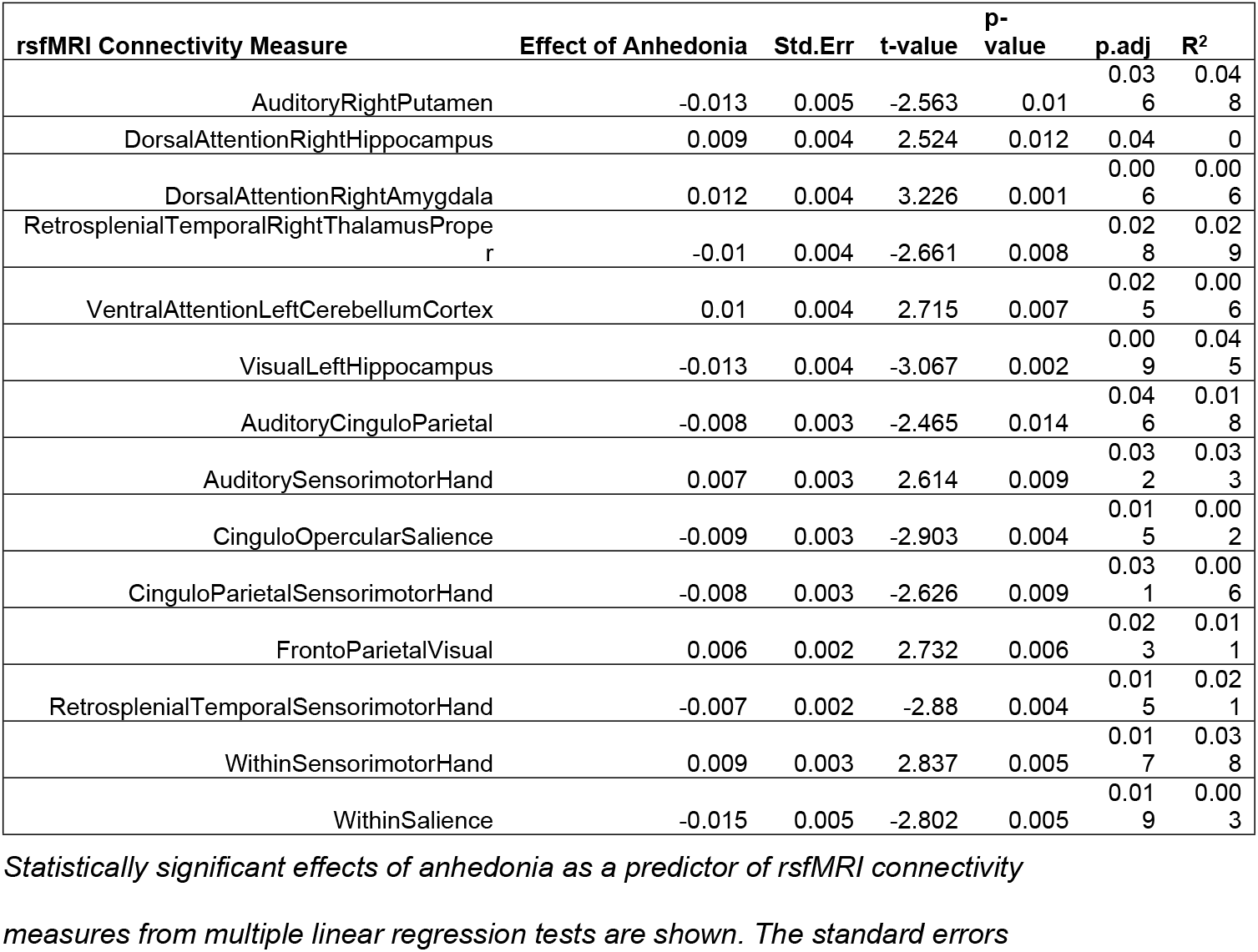
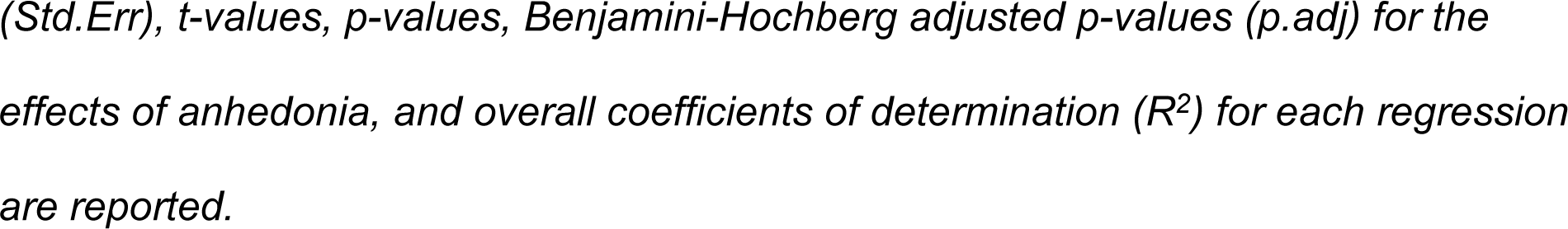
rsfMRI Connectivity Measures with Significant Partial Regression Coefficients for Anhedonia in the ABCD 4.0 Release, Excluding ABCD 1.0 Release, Sub-sample.

In order to maximize our power to detect associations between rsfMRI connectivity measures and anhedonia, we performed multiple linear regression analyses in the full ABCD 4.0 release sample. We found that 16 rsfMRI connectivity measures exhibited significant partial regression coefficients for anhedonia (Table. 6; full table of results found in Supplementary Table 10). Of note, 9 out of these 16 rsfMRI connectivity measures were previously identified to be uniquely associated with anhedonia from our t-tests. Furthermore, the *Retrosplenial Temporal vs. Right Cerebellum Cortex* and *CinguloOpercular vs. Brainstem* connectivity measures were reported to be significantly associated with anhedonia by the previous authors.

**Table 6.** rsfMRI Connectivity Measures with Significant Partial Regression Coefficients for Anhedonia in the Full ABCD 4.0 Release Sample.

| rsfMRI Connectivity Measure | Effect of Anhedonia | Std.Err | t-value | p-value | p.adj | $R^2$ |
| --- | --- | --- | --- | --- | --- | --- |
| RetrosplenialTemporalRightThalamusProper<sup>a</sup> | -0.012 | 0.003 | -3.583 | 0 | 0.001 | 0.029 |
| VentralAttentionLeftCerebellumCortex<sup>a</sup> | 0.009 | 0.003 | 3.161 | 0.002 | 0.006 | 0.008 |
| AuditoryRightPutamen | -0.013 | 0.004 | -3.034 | 0.002 | 0.009 | 0.045 |
| DorsalAttentionRightAmygdala<sup>a</sup> | 0.008 | 0.003 | 2.751 | 0.006 | 0.02 | 0.007 |
| VentralAttentionLeftPutamen | 0.007 | 0.003 | 2.745 | 0.006 | 0.02 | 0.019 |
| CinguloOpercularSalience | -0.007 | 0.003 | -2.667 | 0.008 | 0.025 | 0.002 |
| WithinSensorimotorHand<sup>a</sup> | 0.007 | 0.003 | 2.649 | 0.008 | 0.026 | 0.037 |
| VisualRightPallidum<sup>a</sup> | -0.008 | 0.003 | -2.615 | 0.009 | 0.029 | 0.031 |
| FrontoParietalVisual<sup>a</sup> | 0.005 | 0.002 | 2.609 | 0.009 | 0.029 | 0.012 |
| WithinSalience | -0.011 | 0.005 | -2.449 | 0.014 | 0.04<br>4 | 0.00<br>1 |
| CinguloOpercularBrainStem<sup>a</sup> | -0.007 | 0.003 | -2.437 | 0.015 | 0.04<br>5 | 0.03<br>9 |
| RetrosplenialTemporalSensorimotorHand | -0.005 | 0.002 | -2.422 | 0.015 | 0.04<br>7 | 0.02 |
| VisualRightVentraldc | 0.008 | 0.003 | 2.412 | 0.016 | 0.04<br>8 | 0.00<br>7 |
| RetrosplenialTemporalRightCerebellumCortex<sup>a</sup> | -0.01 | 0.004 | -2.407 | 0.016 | 0.04<br>9 | 0.04<br>1 |
| VentralAttentionLeftCaudate<sup>a</sup> | -0.006 | 0.003 | -2.404 | 0.016 | 0.04<br>9 | 0.02<br>5 |
| SensorimotorHandLeftPallidum | -0.009 | 0.004 | -2.398 | 0.017 | 0.05 | 0.05<br>6 |

Statistically significant effects of anhedonia as a predictor of rsfMRI connectivity measures from multiple linear regression tests are shown. The standard errors (Std.Err), t-values, p-values, Benjamini-Hochberg adjusted p-values (p.adj) for the effects of anhedonia, and overall coefficients of determination ( $R^2$ ) for each regression are reported.
<sup>a</sup> rsfMRI connectivity measures also found to be specifically associated with anhedonia from previously performed t-tests in the full ABCD 4.0 release sample.

Two rsfMRI connectivity measures, the *Auditory vs. Right Putamen* and *Ventral Attention vs. Left Putamen* were previously found to be associated with both depressed mood and anhedonia from our t-tests but now only exhibit significant partial regression coefficients for anhedonia. Interestingly, the *Sensorimotor-Hand vs. Left Pallidum* connectivity measure exhibited significant partial regression coefficients for both anhedonia and depressed mood (Table. 6, Supplementary Table. 10), consistent with previous t-test results, suggesting the presence of significant independent effects of both symptoms on the same rsfMRI connectivity measure.

The *CinguloOpercular vs. Left Amygdala* connectivity measure was also previously associated with both depressed mood and anhedonia, based on their t-tests, but now only exhibit significant partial regression coefficients for depressed mood (Supplementary Table. 10). Similarly, the *Ventral Attention vs. Left Thalamus Proper* connectivity measure was previously associated with both anhedonia and irritability based on t-tests, but now only exhibit significant partial regression coefficients for irritability (Supplementary Table. 10). Only one rsfMRI connectivity measure, *Within Ventral Attention*, exhibited a significant partial regression coefficient for bipolar II disorder (Supplementary Table. 10).

Altogether, while multiple linear regression analyses did not improve replicability between the ABCD study sub-samples, it did allow us to estimate the independent effects of anhedonia on rsfMRI connectivity measures in the full ABCD 4.0 release sample and helped resolve the specificity of the effects of connectivity measures that were previously found to be associated with anhedonia and other psychiatric symptoms.

## Discussion

### Reproduction and replication of previous findings

While we were able to successfully reproduce the previous authors’ findings, we were mostly unable to replicate them using a larger independent subset of the full ABCD 4.0 release sample. Interestingly, a recent study exploring the replicability of brain-behavior association studies using simulations and parametric bootstrapping methods found that relatively small sample sizes (n<500) produced results with significantly inflated effect sizes, low precision, and low replicability and it was only when the sample sizes were increased to the high hundreds or thousands were they able to produce stable effects that were significantly more replicable (22). Thus, it is possible that associations found by the original authors and in our replication analyses using relatively small samples are at risk of being driven by random sample variation.

To maximize statistical power and to reduce inflated associations between rsfMRI connectivity associations with anhedonia, future analyses should be conducted using larger samples, such as in the full ABCD 4.0 release sample. Furthermore, Marek et al., 2022 found that controlling for socio-demographic covariates further reduced effect size inflation. Thus, the results from our multiple linear regression analyses using the full ABCD 4.0 release sample, where we control for socio-demographic covariates, are more likely to represent less-inflated and more replicable findings.

### Specificity of associations

We found depressed mood, irritability, and bipolar II disorder to be significantly comorbid with anhedonia. Using a multiple linear regression approach in the full ABCD 4.0 dataset, we were able to estimate the effects of anhedonia on rsfMRI connectivity measures independent of those comorbid conditions, allowing us to disentangle rsfMRI connectivity measures associated with more than one condition based on t-test results. In doing so, however, the interpretation of anhedonia requires careful consideration and reflection.

As mentioned previously, while anhedonia and depressed mood are core symptoms of major depressive disorders, they are considered distinct processes (15). Alternatively, the hierarchical Taxonomy of Psychopathology (HiTOP) (23), a recently developed dimensional framework for psychopathology, has classified anhedonia as a symptom belonging to two high level *sepctra* of psychopathology; the *internalizing* and *detachment* spectra. Interestingly, low/depressed mood and irritability also fall under the *internalizing* spectrum whereas bipolar II falls under the *thought disorder* spectrum. Thus, when estimating the effects of anhedonia independent of other internalizing and thought disorder related symptoms, we could interpret the remaining effect to emphasize detachment processes. As detachment is a component of the psychosis super-spectrum (24) our findings may represent anhedonic neurocircuitry that may, in part, be related to schizophrenia, schizotypal personality, or other psychotic disorder processes. Further work is required to assess the extent to which differences in rsfMRI connectivity specific to anhedonia better associates with or even predicts dimensional measures of internalizing or detachment related psychopathology.

We included race/ethnicity as a covariate in our multiple linear regression analyses and note they exhibited significant partial regression coefficients for many of the rsfMRI connectivity measures we analyzed. Furthermore, we found there were significantly higher proportions of Black and Hispanic participants in the anhedonia group compared to non-anhedonic controls in the full ABCD 4.0 release sample. Race and ethnicity are social constructs representing complex social and cultural factors (25) deserving careful consideration. Several previous studies have reported significantly higher risk of anhedonia in Black and Hispanic compared to non-Hispanic White adults (26,27) and that these associations may, in part, be accounted for by socioeconomic factors, such as household income and education, as well as other social determinants of health (28), such as disparities in access to healthcare (26). In the ABCD sample, racial discrimination may be an important factor contributing to risk of anhedonia as well as differences in brain-based measures. For example, several recent studies have found that racial discrimination is associated with lower total brain volume (29) and alterations in pre-frontal white matter tracts in adults (30,31). While out of the scope of this study, it will be very important to investigate how social determinants of health and other related factors contribute to differences in health and brain-based outcomes between different racial and ethnic groups during child and adolescent development.

## Limitations

One major limitation of our study was the use of seed-based correlational methods to compute network connectivity measures in functionally-defined networks. As such, these network connectivity measures are averages over large and distributed networks where signals from sub-regions potentially highly associated with anhedonia may be drowned out by signals from sub-regions with low levels of association. Another concern is that the Gordon brain parcellations were produced using a boundary-mapping approach in adult brains (19) so whether they are generalizable to the brains of developing children is important to consider. For example, one study found that the functional topography of connectivity networks does change with age which was predictive of individual differences in executive function (32). One alternative method is to use a decomposition-based method, such as independent components analysis (ICA), to define functional connectivity measures (33). ICA is a data driven approach that extracts components that maximally explain the data and thus, may enhance predictive performance. One study took such an approach and found that using a decomposition-based, compared to a seed-based, extraction of functional networks during a social cognition task achieved significantly greater performance in predicting the degree of social anhedonia in around 70 adolescents/young adults with varying levels of schizotypy (34). Since the raw neuroimaging data from the ABCD data are publicly available, this may be a feasible approach to implement in a future study.

Another limitation in our study is the use of a binary classifier for childhood anhedonia. Several studies have reported greater predictive performance of neuroimaging measures on anhedonia symptom scores (34,35) which suggests that functional neuroimaging measures may be more useful for predicting symptom severity rather than for disorder classification. Thus, it may be more reliable to investigate the associations between functional neuroimaging measures and clinical scales for assessing behavioral problems (such as the Child Behavior Checklist) or neurocognitive performance in individuals with anhedonia.

Finally, the DSM-V definition of anhedonia conflates two distinct reward processes: motivational (interest/wanting) and consummatory (pleasurable/liking) behaviors. These behaviors have been shown to have distinct neurobiological and behavioral components (9,36). We are limited in our study because we do not distinguish between these processes. However, the ABCD study data does include task-based functional neuroimaging of participants completing the monetary incentive delay task, which is able to assess the anticipatory, consummatory, and learning aspects of reward (37). These processes were studied previously by Pornpattananangkul et al., but exceeded the scope of this study. Nevertheless, exploration of brain connectivity specific to each of these reward-based components in subjects with anhedonia may help elucidate the underlying circuitry underlying this complex psychiatric condition.

## Data Availability

All data files are available from the ABCD Study Data Repository from the NIMH Data Archive (https://nda.nih.gov/abcd). The DOI for this NDA Study is 10.15154/1526460.

## Acknowledgements

Data used in the preparation of this article were obtained from the Adolescent Brain Cognitive Development^SM^ (ABCD) Study (https://abcdstudy.org), held in the NIMH Data Archive (NDA). This is a multisite, longitudinal study designed to recruit more than 10,000 children age 9-10 and follow them over 10 years into early adulthood. The ABCD Study® is supported by the National Institutes of Health and additional federal partners under award numbers U01DA041048, U01DA050989, U01DA051016, U01DA041022, U01DA051018, U01DA051037, U01DA050987, U01DA041174, U01DA041106, U01DA041117, U01DA041028, U01DA041134, U01DA050988, U01DA051039, U01DA041156, U01DA041025, U01DA041120, U01DA051038, U01DA041148, U01DA041093, U01DA041089, U24DA041123, U24DA041147. A full list of supporters is available at https://abcdstudy.org/federal-partners.html. A listing of participating sites and a complete listing of the study investigators can be found at https://abcdstudy.org/consortium_members/. ABCD consortium investigators designed and implemented the study and/or provided data but did not necessarily participate in the analysis or writing of this report. This manuscript reflects the views of the authors and may not reflect the opinions or views of the NIH or ABCD consortium investigators.

The ABCD data repository grows and changes over time. The ABCD data used in this report came from http://dx.doi.org/10.15154/1523041.

This manuscript reflects the views of the authors and may not reflect the opinions or views of the NIH or ABCD consortium investigators.

This study received no external funding. Yi Zhou was supported by the department of Psychiatry at Virginia Commonwealth University.

